# Implementation research for advancing health equity: design and competency-based assessment of a short-term implementation research training program focused on health equity in global health

**DOI:** 10.64898/2026.02.03.26345525

**Authors:** Anna Helova, Kevin Owuor, Camryn Durham, Erin Tech, Olakunle Alonge

## Abstract

Implementation research (IR) provides tools to address health inequities and complex global health issues, facilitating real-world health impact at scale. Limited training opportunities exist for applying IR to health inequities. The 2024 Sparkman Center for Global Health Summer Institute (SI) in the School of Public Health, University of Alabama at Birmingham, provided a one-week intensive workshop that introduced concepts, methodologies, and applications of IR in global health settings, explicitly targeting health inequities. This paper describes a competency-based assessment of SI.

We utilized the reliability- and validity-tested Self-evaluation of Implementation Research Competencies and Objective Assessment questionnaires. Analyses included pre/post-SI comparisons of self-assessed knowledge, self-efficacy, and objective knowledge scores; associations between participant characteristics and changes in knowledge/self-efficacy scores; and explored psychometric properties of the objective assessment tool to estimate item difficulty and discrimination parameters. Open-ended questions were analyzed thematically to reveal participant experiences, perceived relevance of the training, and suggestions for future improvement.

SI was completed by 28 participants from 12 countries working on various global health topics. Equity considerations within projects focused on social/structural barriers, differentiating outcomes, and strategies to target disadvantaged groups within IR. Significant improvements were observed across IR domains, with the mean self-assessed knowledge increasing from 57.67% [Confidence interval (CI):43.78; 71.55] to 87.68% (CI: 76.96;98.41), p=0.003, and the largest improvements in competencies related to stakeholder engagement, scientific inquiry, and the application of IR strategies to real-world problems. Objective assessment scores increased from 65.35% (CI: 61.28;69.41) to 68.18% (CI: 64.38;71.97), p=0.324). Participants reported high satisfaction with SI’s structure, content, and delivery, highlighting its practical focus on stakeholder engagement, real-world case studies, mentorship, networking, calling for continued mentorship engagement and further training. This evaluation will be used to improve the impact of IR training in global health settings, emphasizing importance of hands-on application of IR concepts in tackling health inequities.

## INTRODUCTION

Implementation Research (IR) addresses questions pertaining to implementation of evidence-supported health interventions (ESI) to achieve population health impact.[1] IR has been criticized for not paying sufficient attention to issues of health equity[2], that is, “everyone having a fair and just opportunity to be as healthy as possible”[3] – which is much needed to achieve widespread population health impact in global health settings.[2, 4, 5] Similarly, while there are several degrees and training programs in implementation research relevant to global health, there are very few that explicitly target issues of health inequities contextualized to global health settings.[6] Global health settings include resource-limited settings where the world’s most vulnerable and least disadvantaged populations live, and are not restricted to low- and middle-income countries (LMICs).[7, 8]

Beyond the lack of a specific focus on equity issues, trainings in implementation research are inaccessible or not as impactful for many practitioners and researchers due to several reasons: inaccessibility of long-term, in-person academic programs due to costs and time and other logistical reasons, focus on basic IR principles with no real-life application, limited content/geographic location focus, limited contextual validity of content for a diversified audience, and lack of clear linkage into a career pipeline. These reasons disproportionately affect all trainees in global health settings.[9–11] Additionally, while competency-based training in global health is considered highly valuable,[12, 13] many IR training programs in global health settings fail to clarify the competencies they teach and may not cover advanced competencies needed to tackle health inequities.[14] Moreover, there are limited standardized and validated methodologies for evaluating the impact of competency-based training programs in IR, particularly in global health settings.[15]

To address some of these gaps, the Sparkman Center for Global Health (SCGH) Summer Institute offers an annual one-week intensive workshop that introduces implementation research (IR) in global health settings and provides competency-based training in applying IR concepts and methods to address health inequities in global health research and practice. The first Institute was a one-week residential workshop held in Birmingham, Alabama, in July 2024. The SI aimed to equip participants to: (1) articulate key principles of IR; (2) distinguish how IR differs in global health from non-global health settings; (3) apply relevant IR approaches to global health and practice; and (4) operationalize IR to address health inequity in a global setting. The training format included a mix of lectures, moderated discussions, breakout groups, and cultural experiences in Birmingham, Alabama, given the city’s historical role in confronting issues of racial inequities in the United States. The week concluded with case presentations from all trainees and structured feedback to improve their proposals.

The objective of this paper is to describe methods and findings from a competency-based assessment of the 2024 SCGH Summer Institute in Implementation Research in Global Health with a focus on health equity. The paper follows methods described for competency-based assessment of implementation research training programs in low- and middle-income countries.[15] This paper aims to provide insights into the impact of competency-based IR training programs for researchers and practitioners, and how these programs can be designed to address advanced competencies in IR, including tackling health equity and other gaps identified in the literature. The findings from this paper will contribute to methods and tools for competency-based assessment of short-term IR training and will be used to improve future iterations of the SCGH Summer Institute and similar training programs in global health settings.

## METHODS

### Setting and recruitment

The SCGH 2024 Summer Institute was held at the UAB School of Public Health (SOPH) in the heart of Birmingham, Alabama, a city established in 1871 with a rich civil rights history in the United States. Lectures and discussions were organized in SOPH classrooms equipped with high-tech facilities. Participants were accommodated in UAB dormitories, hotels, or on their own, based on their preferences and budget, all within walking distance of the SOPH facilities.

Professionals working in global health with advanced degrees were the primary audience, including post-doctoral students. A recruitment and communication plan was developed to attract this audience and included communications distributed domestically and internationally through various global health media, newsletters, and networks, including the NIH Fogarty Launch the Future Leaders of Global Health (LAUNCH) Network, the Consortium of Universities for Global Health (CUGH), and the American Public Health Association. Tiered registration pricing and scholarship availability facilitated access to a diverse pool of participants.

### Approach

The Institute was designed and curriculum developed by the senior author, OA, drawing on his experience and expertise teaching and conducting research in implementation science and health equity in global health settings, with the support of other IR experts and facilitators (Table 1), including IR experts with expertise in implementation science and equity research in diverse geopolitical settings, educational background, methodological approaches, and topics. Table 1 provides details on the topics covered, the competencies addressed, and the pedagogical strategies applied to address each topic. Topics covered included IR in global health; IS theories, models, and frameworks relevant to global health; application of selected IR methods to global health research and practice; and how to leverage IR for health equity in global health. The workshop combined four pedagogical strategies, including high-level lectures, case examples, a problem-solving clinic, and visit to the Birmingham Civil Rights Institute (BCRI) to provide contexts for discussion on equity.

**Table 1.** Summer Institute sessions, topics, competencies, and pedagogical strategies.

| Topics covered | Competencies Addressed | Pedagogical Strategies Applied |
| --- | --- | --- |
| (1) Implementation research in global health – what is it and how to do it<br>(2) Implementation of science theories, models, and frameworks relevant to global health (resource-limited settings)<br>(3) Application of selected IR methods to global health research and practice<br>(4) How to leverage IR for health equity in global health. | (1) Articulate key principles of IR<br>(2) Distinguish how IR differs in global health from non-global health settings<br>(3) Apply relevant IR approaches to global health and practice<br>(4) Operationalize IR to address health inequity in a global setting. | (1) High-level lectures introducing IR in global health, and relevant IR concepts and approaches for addressing health inequities in global health research and practice<br>(2) Case examples demonstrating the application of concepts and approaches from the lectures to real-world global health research and practice cases<br>(3) Problem-solving clinic working with participants to apply IR concepts and approaches to their own global health research, practice, and/or policy cases to address health inequities.<br>(4) Cultural experiences in Birmingham, including a visit to the Civil Rights Institute in Birmingham, Alabama. The BCRI is a cultural and educational research center that promotes a comprehensive understanding of the significance of civil rights developments in Birmingham, Alabama. |

| Detailed Schedule | Morning session | Afternoon session |
| --- | --- | --- |
| **DAY 1:** | **Lecture 1:** Implementation research in global health – what is it and how to do it? | **Workshop 1:** Application of key IR principles and building blocks to draft case examples & **Group activities 1:** Participants present cases and progress to each other in small groups |
| **DAY 2:** | **Lecture 2a:** Application of selected IR methods to global health research and practice objectives (Quantitative) & **Lecture 2b:** Application of selected IR methods to global health research and practice objectives (Qualitative and mixed methods) | **Workshop 2:** Workshop 2 (Application of relevant IR research methods to draft case examples) & **Group activities 2:** Participants present cases and progress to each other in small groups |
| **DAY 3:** | **Lecture 3:** Implementation science (IS) theories, models, and frameworks (TMFs) relevant to global health | **Workshop 3:** Application of IS TMF to draft case example & **Group activities 3:** Participants present cases and progress to each other in small groups |
| **DAY 4:** | **Lecture 4:** IS theories, models, and frameworks relevant for addressing health inequities in global health settings | **Group activities 4:** Application of equity-focused IR framework and worksheets to draft case examples. Participants present cases and progress to each other in small groups |
| **DAY 5:** | **Workshop 4:** How to leverage IR for equity in global health & **individual activities** | **Individual activities & Group activities 5:** Application of equity-focused IR framework and worksheets to draft case examples. Participants present cases and progress to each other in small groups |
| **DAY 6:** | **Summary, overview, and assessments & presentation of cases** with faculty and peer feedback | **Presentation of cases** with faculty and peer feedback & completion certificates **award ceremony** |

The SI concluded with case presentations of proposals for research/practice-related projects from all trainees developed before and during the Institute, through knowledge gain, peer-to-peer engagements, and faculty-led discussions. To guide systematic identification and assessment of equity considerations throughout their project’s lifecycle, equity analysis frameworks and worksheet were developed, and participants were guided on how to apply them.[2] These frameworks and worksheets make equity considerations explicit in IR design and reporting, ensuring equity across different populations, inclusion of vulnerable populations, and integration of structural and contextual factors as part of IR research/practice agenda.[16] Most participants stayed in the same location, further facilitating ongoing peer discussions.

A welcome packet was developed and distributed prior to the SI to provide logistical details. A secure folder using the UAB Box platform was established to share lectures, readings, and other resources with participants prior to and during the workshop. Daily information emails were distributed to highlight the daily schedule and logistical details. The SCGH continues to engage with participants to build upon SI training and projects and recruit participants for subsequent SIs.

### Data collection and assessment tools

We utilized the competency-based assessment data collection tools for IR training programs in low- and middle-income countries (LMIC), developed by Alonge, et al.[15], which in turn are based on a framework of IR core competencies in LMICs.[14] Briefly, the data collection tools included pre- and post-SI assessments, utilizing: (a)*the Self-evaluation of Implementation Research Competencies* and b) *the Objective Assessment questionnaires*.

The *Self-evaluation questionnaires* consist of (1) a 16-item self-assessment scale for IR knowledge evaluated across six IR themes (working with stakeholders, scientific inquiry, implementation strategies, resources for IR competencies, communication and advocacy, and cross-cutting themes, including contexts and ethical considerations); (2) a 16-item self-assessment scale for IR self-efficacy to assess confidence of application of six IR themes included in the IR knowledge scale; (3) ten questions assessing prior experience in application or training others in IR competencies; and (4) a post-SI survey comprising of seven structured questions assessing participants satisfaction and perceived quality of the SI training – all rated on a Likert scale from 1=strongly disagree to 7=strongly agree. Moderate to high knowledge and/or confidence across IR competencies was defined as a rating of 5 or higher on the Likert scale. Similarly, moderate to high satisfaction was defined as a rating of 5 or higher.

*The Objective Assessment* consists of 40 true-false statements assessing knowledge across six IR themes focused on IR principles, concepts, and methodologies. A prepared answer key was used to determine the correct response to each question. Both the self-evaluation and objective assessment tools report high validity and reliability based on exploratory factor analyses and a one-parameter logistic model study to establish construct validity and internal consistency of the tools.[15]

For the SCGH SI, the tools were additionally adapted as follows: The pre-SI questionnaire assessing IR competencies included questions about the participant’s number of years working in global health, current professional position, and an open-ended question about any previous IR courses taken, and the post-SI assessment included optional open-ended questions to gather feedback on the quality and content of the training.

All assessments were disseminated to the participants via a password-protected Qualtrics dashboard. Contact information provided during the SI registration was used to generate a unique identifier for accessing Qualtrics surveys. Each participant received an email containing this unique identifier, not related to or derived from information that identifies the participant. The pre-Institute assessments were sent via the Qualtrics platform a week before the SI. The post-SI assessments were sent at the conclusion of the SI. Automatic reminders during the week prior to SI and three weeks after the SI were sent to ensure high response rates. Within the Qualtrics platform, all questions were set to require a response to avoid omissions by mistake and to maintain the reliability and validity of the instruments. Participants could select “Not Applicable, N/A” if a question was not relevant. Safeguards were put in place to protect the privacy of study participants, and all identifiable data was scrubbed from the dataset before analysis. Data missingness was examined to identify any patterns or potential bias. There were no material benefits for the participants in this study. All data have been stored in the UAB Box. This study was exempt as non-human subject research by the UAB research ethics board. Ethical approval for this study has been obtained from the Institutional Review Board at the University of Alabama at Birmingham, protocol number IRB-300013258. The data were accessed for research purposes on November 5, 2024. Authors have not had access to information that could identify individual participants during or after data collection.

### Evaluation design and data analysis

A pre- and post-quantitative evaluation design was used to assess participant knowledge and self-efficacy. Quantitative data collected from pre- and post-SI assessments were analyzed using Stata 18 (StataCorp, College Station, TX, USA). Descriptive statistics, including means and standard deviations for continuous variables and frequencies and percentages for categorical variables, were used to summarize participant demographic characteristics and baseline knowledge and self-efficacy in IR competencies. Self-assessed knowledge, objectively assessed knowledge, and self-efficacy scores were compared before and after the SI using t-tests for normally distributed variables and ranksum tests when normality assumptions were not met. In addition, the correlation of self-assessed and objectively assessed knowledge scores was estimated before and after the SI using Pearson’s correlation. A paired analysis was not feasible given an error in capturing unique identifiers. Associations between participant characteristics and changes in knowledge or self-efficacy scores were assessed using chi-square or Fisher’s exact tests for categorical variables and independent samples t-tests or Mann–Whitney U tests for continuous variables. To evaluate the psychometric properties of the objective assessment tool in the context of the SCGH SI, a one-parameter logistic model (Rasch model) was applied separately to pre- and post-assessment data to estimate item difficulty and discrimination parameters. Stability in item difficulty across assessments was used to assess the construct validity of the tool. Missing data were minimal (<5%) and analyses were based on available cases without imputation. Additionally, responses to open-ended survey questions were analyzed thematically [17, 18], allowing for the identification of recurring themes related to participant experiences, perceived relevance of the training, and suggestions for future improvement.

## RESULTS

Overall, 28 participants from 12 countries attended the Sparkman Center for Global Health Summer Institute in July 2024 (Table 2). Women constituted 46.4% (n=13) of participants. Over 40% of participants reported 5 or more years of global health experience at baseline, and approximately half had prior exposure to IR coursework, mainly at the introductory level.

**Table 2.** Sociodemographic and professional characteristics of Summer Institute participants.

| IR course before workshop (N=28) | n (%) |
| --- | --- |
| *Gender:* |  |
| Women | 13 (46.4) |
| Men | 15 (53.6) |
| *Region:* |  |
| United States | 8 (28.6) |
| Africa | 14 (50.0) |
| Asia | 5 (17.8) |
| Latin America and the Caribbean | 1 (3.6) |
| *Number of years working in global health:* |  |
| Less than 1 year | 6 (24.0) |
| 1-4 years | 9 (36.0) |
| 5-10 years | 7 (28.0) |
| More than 10 years | 3 (12.0) |
| *Current Position:* |  |
| Trainee/Student | 8 (32.0) |
| Academic professional (e.g., faculty) | 8 (32.0) |
| Non-academic professional (e.g., program managers) | 9 (36.0) |

During the SI, participants developed their projects. The topical focus, geographic area, IR theories, models, and frameworks (TMFs) utilized and equity considerations for those projects are listed in Box 1. Most projects were set in LMICs, particularly in the African Region and Southeast Asia. Topically, most projects focused on maternal and child health, infectious diseases (HIV in particular), non-communicable diseases including cardiovascular issues, various types of cancer, and mental health. Most trainees used the Consolidated Framework for Implementation Research (CFIR)[19], Exploration, Preparation, Intervention, Sustainment (EPIS)[20], and Reach, Effectiveness, Adoption, Implementation, Maintenance (RE-AIM) frameworks[21], often combined with other theoretical models/frameworks, e.g., Health Beliefs or the Socioecological Model. Equity considerations addressed vulnerable populations in low-resource settings globally, particularly those residing in rural communities.

#### Box 1. Focus of the implementation of Science Research Projects of the Summer Institute participants

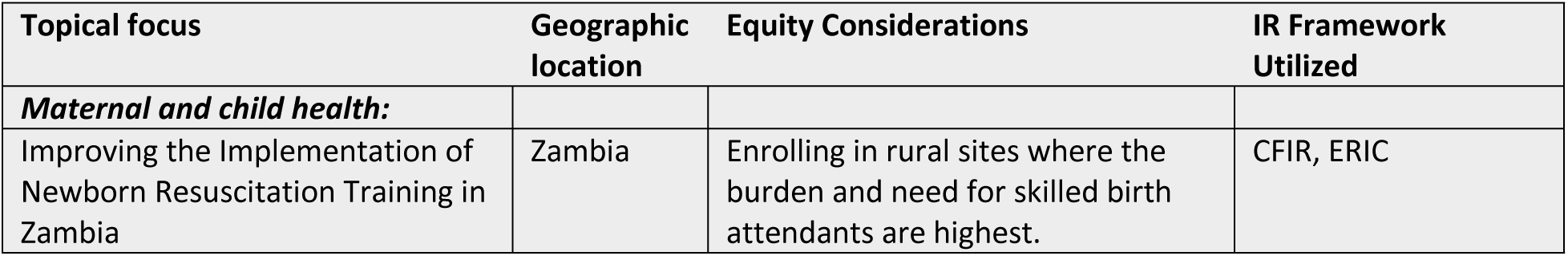
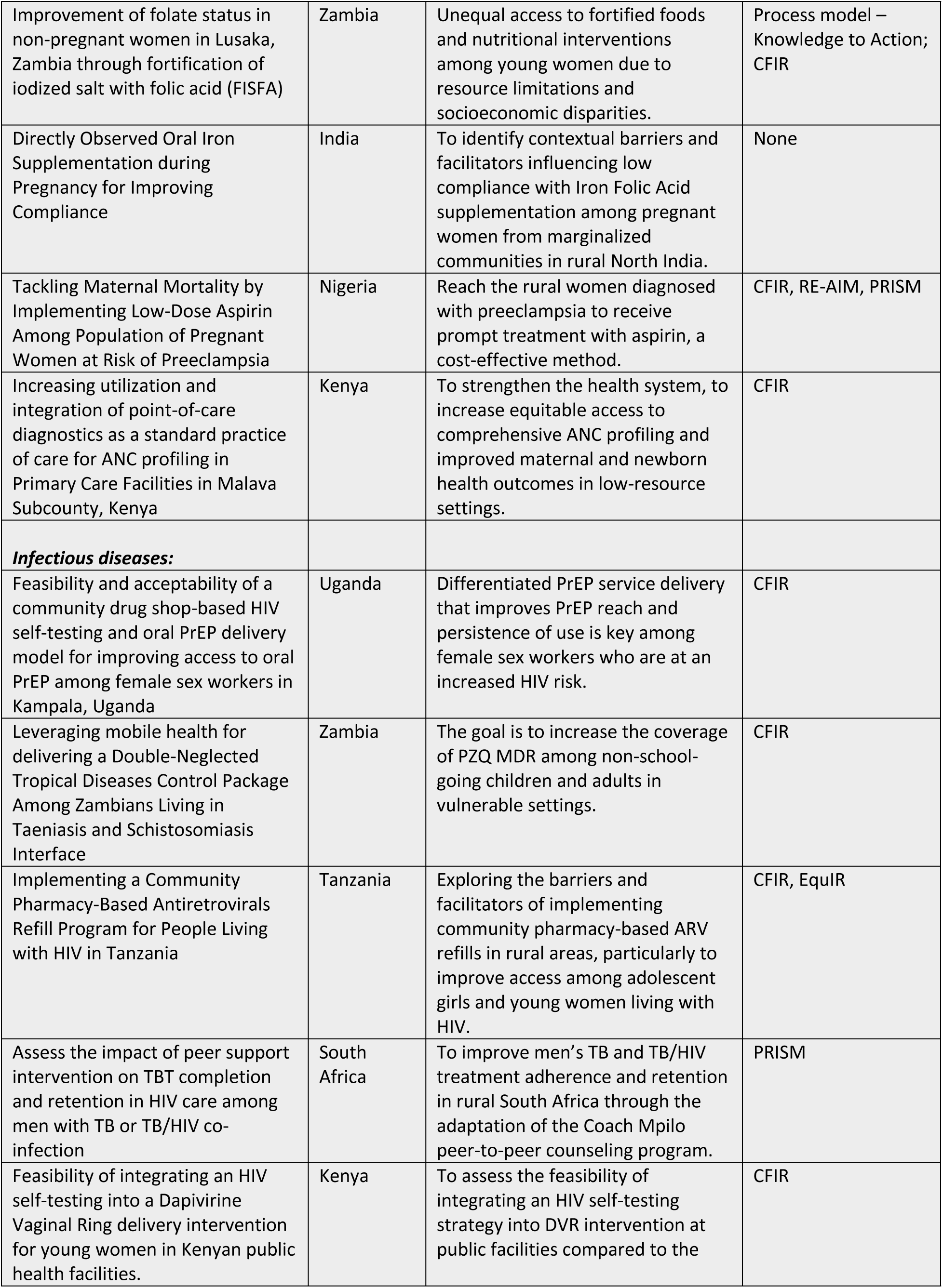
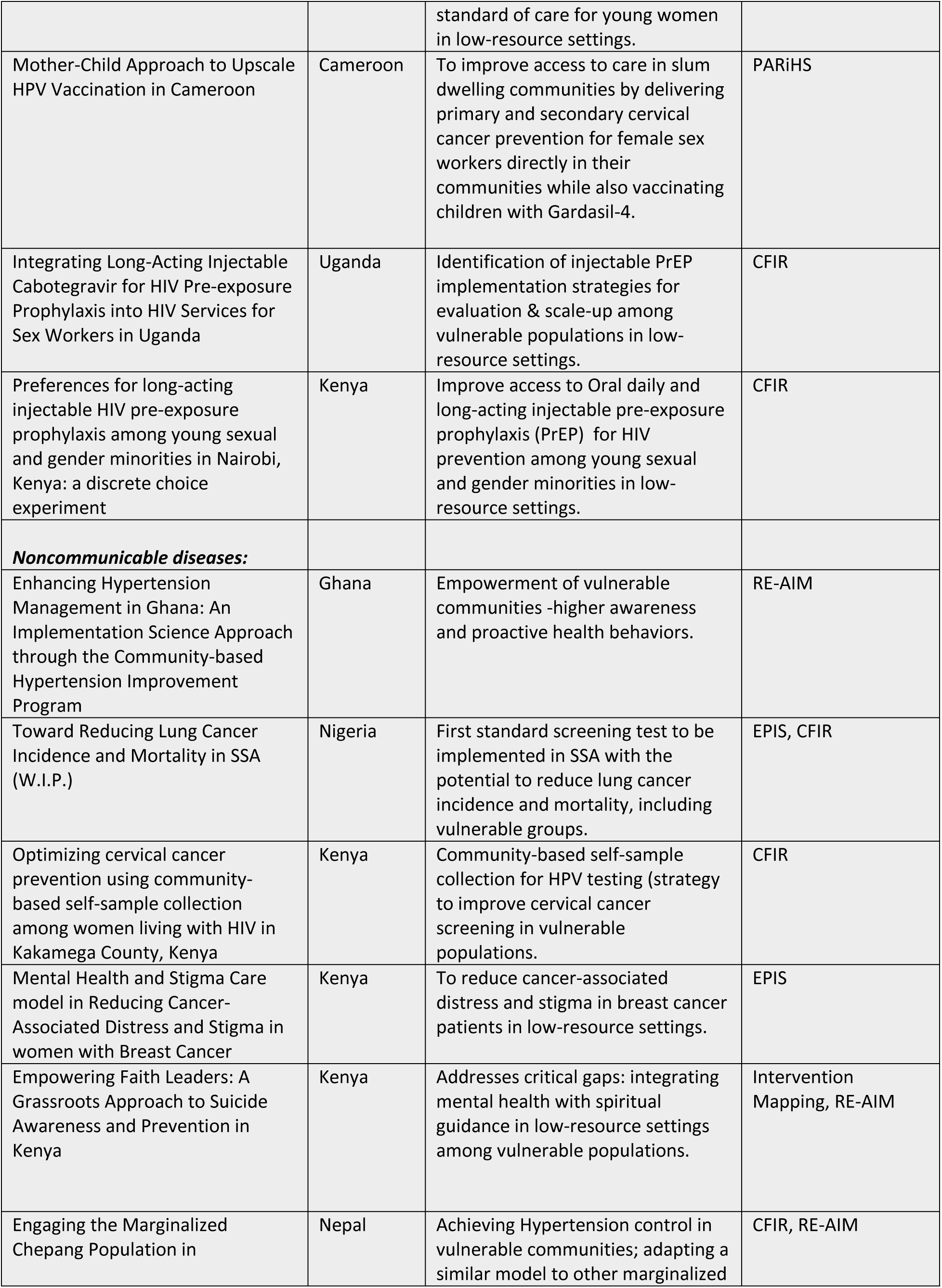
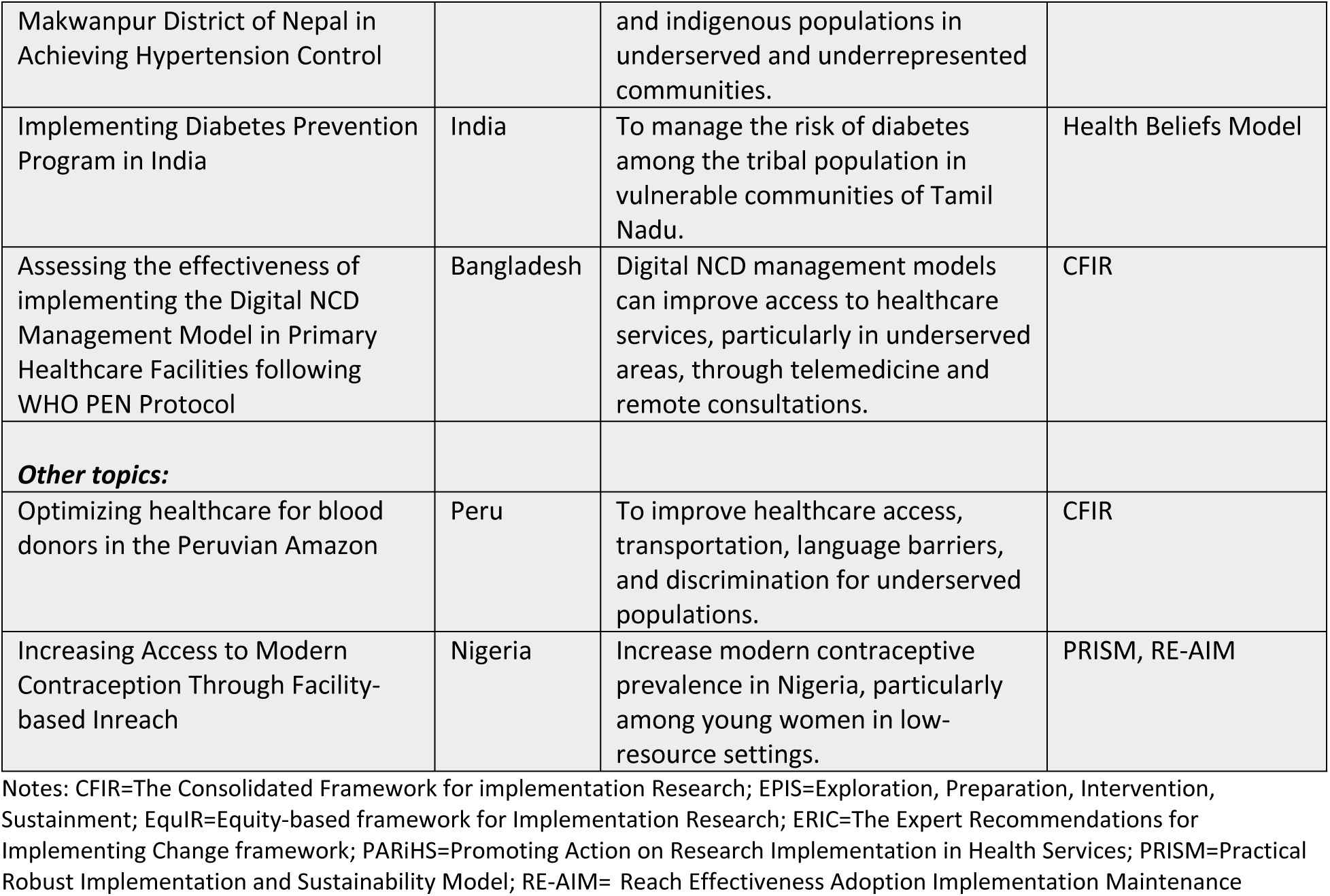

### Self-evaluation of Implementation Research Competencies

Pre-SI self-assessment data revealed that over half of participants reported a moderate confidence in engaging stakeholders (Table 3). However, confidence levels were notably lower for competencies related to conducting IR robustly and rigorously. Participants showed marked improvements in both self-assessed IR knowledge and self-efficacy across all six thematic domains after the IR course (Table 3). Statistically significant gains (p<0.05 for all items) were observed in most competencies of IR knowledge: scientific inquiry, implementation strategies, resources for IR, and communication and advocacy. Similarly, statistically significant gains (p<0.05) were observed in most competencies of self-efficacy: scientific inquiry, implementation strategies, and communication and advocacy. Post-SI assessment data showed a significant overall improvement observed across all self-assessed domains (Table 4) with the mean self-assessed knowledge and confidence increasing by 30.01 percentage points [from 57.67% (95% ci: 43.78; 71.55) to 87.68% (95% ci: 76.96; 98.41), p=0.003].

**Table 3.**
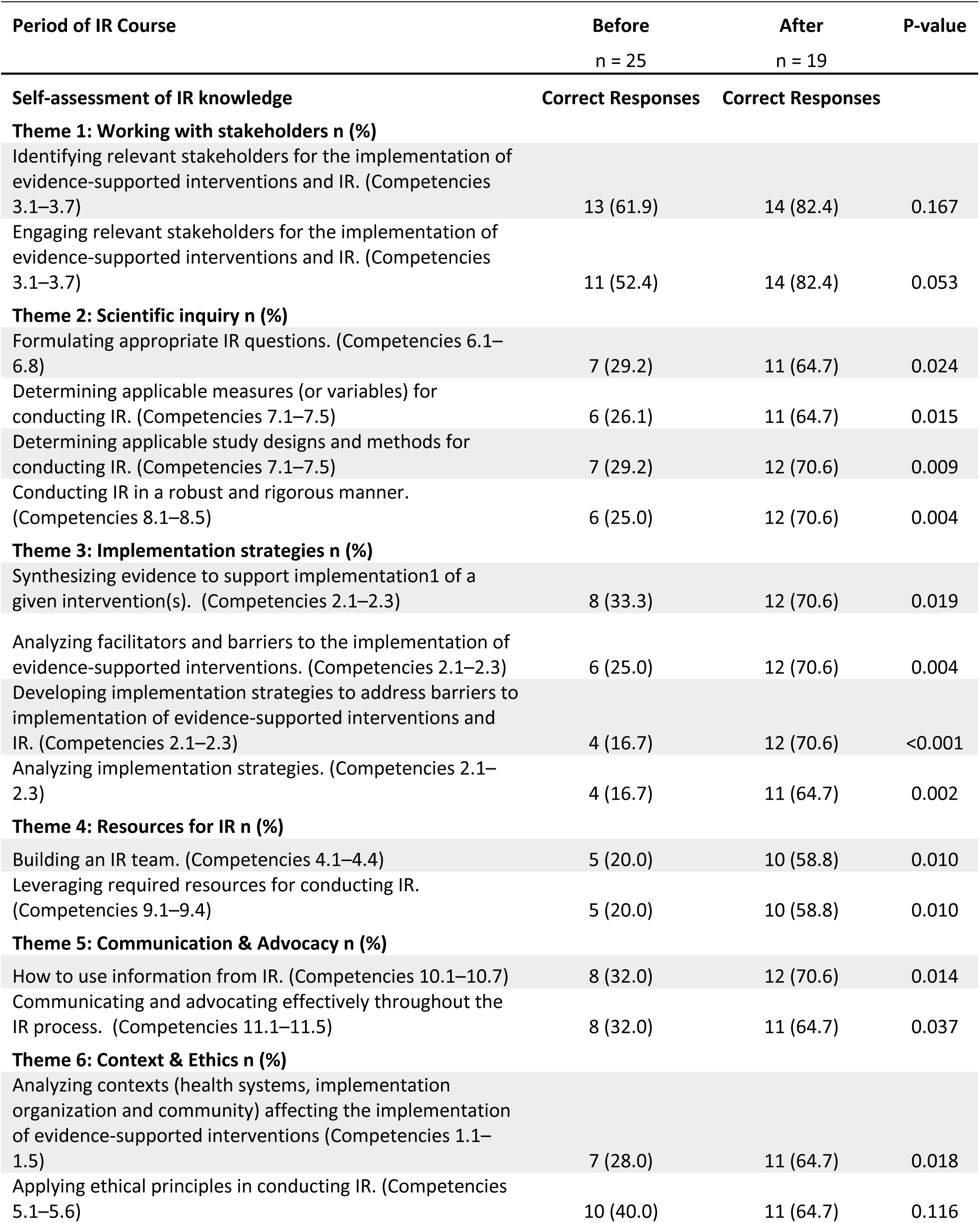
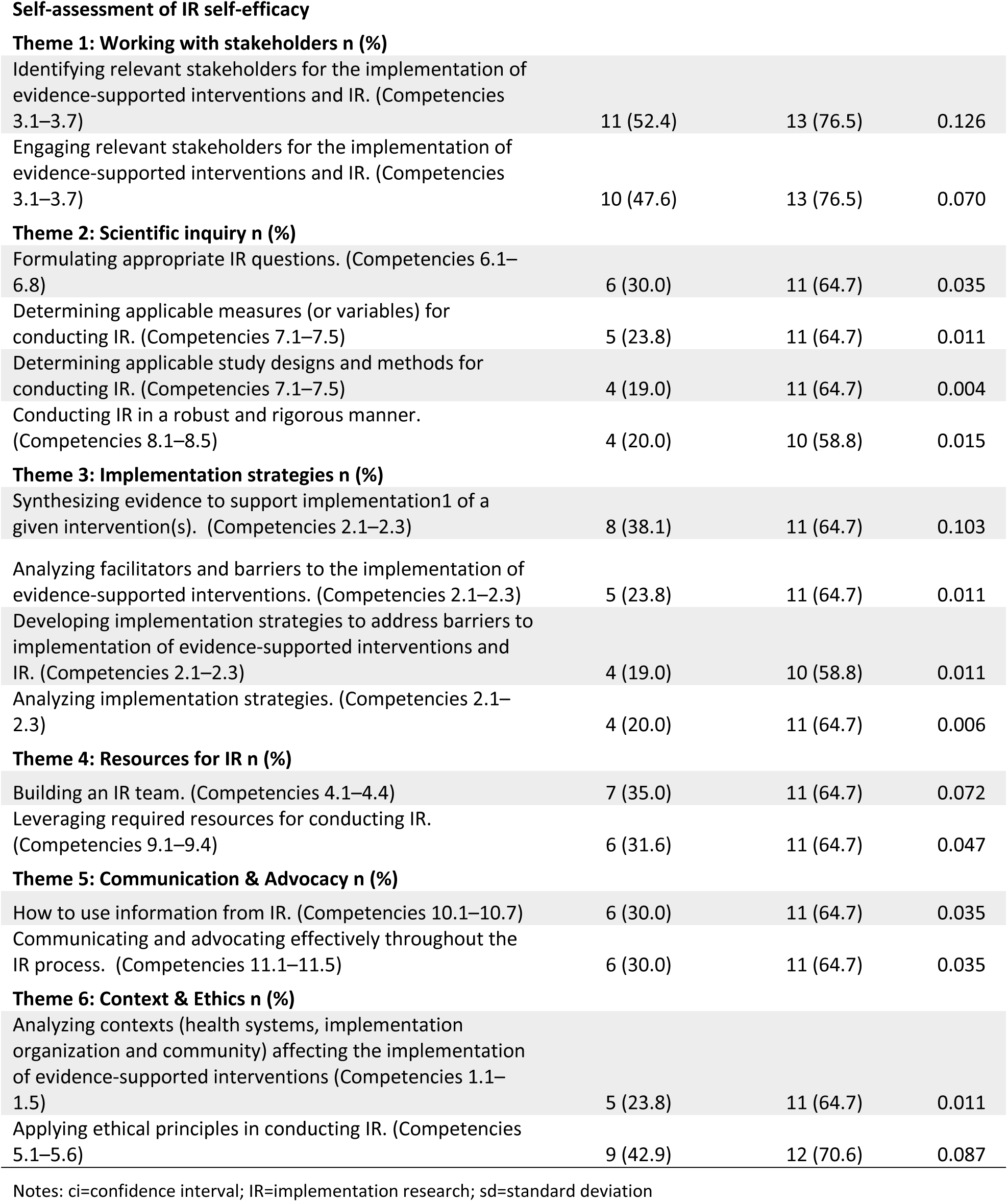
Self-evaluation of Implementation Research Competencies.

| Period of IR Course | Before<br>n = 25 | After<br>n = 19 | P-value |
| --- | --- | --- | --- |
| **Self-assessment of IR knowledge** | **Correct Responses** | **Correct Responses** |  |
| **Theme 1: Working with stakeholders n (%)** |  |  |  |
| Identifying relevant stakeholders for the implementation of evidence-supported interventions and IR. (Competencies 3.1–3.7) | 13 (61.9) | 14 (82.4) | 0.167 |
| Engaging relevant stakeholders for the implementation of evidence-supported interventions and IR. (Competencies 3.1–3.7) | 11 (52.4) | 14 (82.4) | 0.053 |
| **Theme 2: Scientific inquiry n (%)** |  |  |  |
| Formulating appropriate IR questions. (Competencies 6.1–6.8) | 7 (29.2) | 11 (64.7) | 0.024 |
| Determining applicable measures (or variables) for conducting IR. (Competencies 7.1–7.5) | 6 (26.1) | 11 (64.7) | 0.015 |
| Determining applicable study designs and methods for conducting IR. (Competencies 7.1–7.5) | 7 (29.2) | 12 (70.6) | 0.009 |
| Conducting IR in a robust and rigorous manner. (Competencies 8.1–8.5) | 6 (25.0) | 12 (70.6) | 0.004 |
| **Theme 3: Implementation strategies n (%)** |  |  |  |
| Synthesizing evidence to support implementation1 of a given intervention(s). (Competencies 2.1–2.3) | 8 (33.3) | 12 (70.6) | 0.019 |
| Analyzing facilitators and barriers to the implementation of evidence-supported interventions. (Competencies 2.1–2.3) | 6 (25.0) | 12 (70.6) | 0.004 |
| Developing implementation strategies to address barriers to implementation of evidence-supported interventions and IR. (Competencies 2.1–2.3) | 4 (16.7) | 12 (70.6) | <0.001 |
| Analyzing implementation strategies. (Competencies 2.1–2.3) | 4 (16.7) | 11 (64.7) | 0.002 |
| **Theme 4: Resources for IR n (%)** |  |  |  |
| Building an IR team. (Competencies 4.1–4.4) | 5 (20.0) | 10 (58.8) | 0.010 |
| Leveraging required resources for conducting IR. (Competencies 9.1–9.4) | 5 (20.0) | 10 (58.8) | 0.010 |
| **Theme 5: Communication & Advocacy n (%)** |  |  |  |
| How to use information from IR. (Competencies 10.1–10.7) | 8 (32.0) | 12 (70.6) | 0.014 |
| Communicating and advocating effectively throughout the IR process. (Competencies 11.1–11.5) | 8 (32.0) | 11 (64.7) | 0.037 |
| **Theme 6: Context & Ethics n (%)** |  |  |  |
| Analyzing contexts (health systems, implementation organization and community) affecting the implementation of evidence-supported interventions (Competencies 1.1–1.5) | 7 (28.0) | 11 (64.7) | 0.018 |
| Applying ethical principles in conducting IR. (Competencies 5.1–5.6) | 10 (40.0) | 11 (64.7) | 0.116 |
| Theme 1: Working with stakeholders n (%) |  |  |  |
| Identifying relevant stakeholders for the implementation of evidence-supported interventions and IR. (Competencies 3.1–3.7) | 11 (52.4) | 13 (76.5) | 0.126 |
| Engaging relevant stakeholders for the implementation of evidence-supported interventions and IR. (Competencies 3.1–3.7) | 10 (47.6) | 13 (76.5) | 0.070 |
| Theme 2: Scientific inquiry n (%) |  |  |  |
| Formulating appropriate IR questions. (Competencies 6.1–6.8) | 6 (30.0) | 11 (64.7) | 0.035 |
| Determining applicable measures (or variables) for conducting IR. (Competencies 7.1–7.5) | 5 (23.8) | 11 (64.7) | 0.011 |
| Determining applicable study designs and methods for conducting IR. (Competencies 7.1–7.5) | 4 (19.0) | 11 (64.7) | 0.004 |
| Conducting IR in a robust and rigorous manner. (Competencies 8.1–8.5) | 4 (20.0) | 10 (58.8) | 0.015 |
| Theme 3: Implementation strategies n (%) |  |  |  |
| Synthesizing evidence to support implementation of a given intervention(s). (Competencies 2.1–2.3) | 8 (38.1) | 11 (64.7) | 0.103 |
| Analyzing facilitators and barriers to the implementation of evidence-supported interventions. (Competencies 2.1–2.3) | 5 (23.8) | 11 (64.7) | 0.011 |
| Developing implementation strategies to address barriers to implementation of evidence-supported interventions and IR. (Competencies 2.1–2.3) | 4 (19.0) | 10 (58.8) | 0.011 |
| Analyzing implementation strategies. (Competencies 2.1–2.3) | 4 (20.0) | 11 (64.7) | 0.006 |
| Theme 4: Resources for IR n (%) |  |  |  |
| Building an IR team. (Competencies 4.1–4.4) | 7 (35.0) | 11 (64.7) | 0.072 |
| Leveraging required resources for conducting IR. (Competencies 9.1–9.4) | 6 (31.6) | 11 (64.7) | 0.047 |
| Theme 5: Communication & Advocacy n (%) |  |  |  |
| How to use information from IR. (Competencies 10.1–10.7) | 6 (30.0) | 11 (64.7) | 0.035 |
| Communicating and advocating effectively throughout the IR process. (Competencies 11.1–11.5) | 6 (30.0) | 11 (64.7) | 0.035 |
| Theme 6: Context & Ethics n (%) |  |  |  |
| Analyzing contexts (health systems, implementation organization and community) affecting the implementation of evidence-supported interventions (Competencies 1.1–1.5) | 5 (23.8) | 11 (64.7) | 0.011 |
| Applying ethical principles in conducting IR. (Competencies 5.1–5.6) | 9 (42.9) | 12 (70.6) | 0.087 |

Notes: ci=confidence interval; IR=implementation research; sd=standard deviation

**Table 4.** Participants’ Implementation Research (IR) Activities.

| Period of IR Course | Before | After | P-value |
| --- | --- | --- | --- |

|  | n = 25 | n = 19 |  |
| --- | --- | --- | --- |
| **IR activities** | **Correct Responses** | **Correct Responses** |  |
| **Applying IR n (%)** |  |  |  |
| Using IR theories, models and frameworks in a new or existing project | 5 (26.3) | 11 (64.7) | 0.021 |
| Using IR measures, methods or study design in a new or existing project | 3 (15.8) | 14 (82.4) | <0.001 |
| Using systematic approaches in IR to identify and work with different stakeholders in a new or existing project | 3 (15.8) | 12 (70.6) | 0.001 |
| Using systematic approaches in IR to disseminate and communicate research findings to different stakeholders. | 2 (10.5) | 10 (58.8) | 0.002 |
| Using IR materials (e.g., slides, activity sheets, resources) in your implementation project. | 3 (15.8) | 12 (70.6) | 0.001 |
| **Training others in IR n (%)** |  |  |  |
| Sharing IR course materials (e.g., slides, activity sheets, resources) within your network or with members from external organizations. | 2 (10.5) | 10 (58.8) | 0.002 |
| Training other people on how to apply IR theories, models and frameworks. | 1 (5.3) | 6 (35.3) | 0.023 |
| Training other people on how to apply IR measures, methods or study design. | 0 (0.0) | 6 (35.3) | 0.006 |
| Training other people on how to use systematic approaches in IR to identify and work with different stakeholders in a new or existing project. | 1 (5.3) | 7 (41.2) | 0.010 |
| Training other on how to use systematic approaches in IR to disseminate and communication findings to different stakeholders. | 1 (5.3) | 7 (41.2) | 0.010 |
| **Implementation Quality and Participant Satisfaction n (%)** |  |  |  |
| I was extremely satisfied with the session readings and resources. | N/A | 17 (94.4) | N/A |
| Overall, I was satisfied with the presentations. | N/A | 17 (94.4) | N/A |
| Overall, I was satisfied with how the content applies to my work. | N/A | 17 (94.4) | N/A |
| I was satisfied with the session activities. | N/A | 17 (94.4) | N/A |
| I was satisfied with the format of the session (presentation, group activity etc.) | N/A | 17 (94.4) | N/A |
| I was satisfied with the content of the sessions. | N/A | 17 (94.4) | N/A |
| Overall, I thought the course was implemented with high quality. | N/A | 17 (94.4) | N/A |
| **Summary Scores** |  |  |  |
| Percentage Total Score, mean (sd) | 57.67 (35.42) | 87.68 (23.86) | 0.003 |
| Percentage Total Score, mean (95% ci) | 57.67 (43.78; 71.55) | 87.68 (76.96; 98.41) | 0.003 |
| Total Score, mean (sd) | 23.32 (16.33) | 38.37 (16.18) | 0.004 |
| Total Score, mean (95% ci) | 23.32 (16.92; 29.72) | 38.37 (31.09; 45.65) | 0.004 |

Notes: ci=confidence interval; IR=implementation research; sd=standard deviation

### Objective Assessment questionnaire

Objective assessment scores also demonstrated improvement. The percentage mean score on the 40-item objective test increased by 2.83 percentage points [from 65.35% (95% ci: 61.28; 69.41) before to 68.18% (95% ci: 64.38; 71.97) after the IR workshop, p = 0.324] [Table 5]. Item Response Theory (IRT) analysis showed that item difficulty parameters remained stable between the pre- and post-assessments, supporting the structural validity of the objective tool [Figure 1(a) and Figure 1(b)]. However, the discrimination parameter decreased from 0.79 before the IR workshop to 0.63 after the workshop, suggesting reduced ability of the tool to discriminate between high and low-ability participants after training. Overall, participants reported a high level of satisfaction with the SI, with over 94% expressing that the course was implemented with high quality and was directly applicable to their professional work. At baseline, the correlation between self-assessed knowledge and objectively assessed knowledge was low (r=0.05, p=0.804), indicating limited agreement between perceived and actual competence. Post-SI feedback from participants highlighted high satisfaction with the IR workshop structure, content, and delivery. Common suggestions included allocating more time for practical exercises, providing ongoing mentorship opportunities, and offering additional follow-up sessions to reinforce learning and support application in participant work contexts. Participants highlighted the practical focus on stakeholder engagement and the use of real-world case studies.

**Fig 1.**
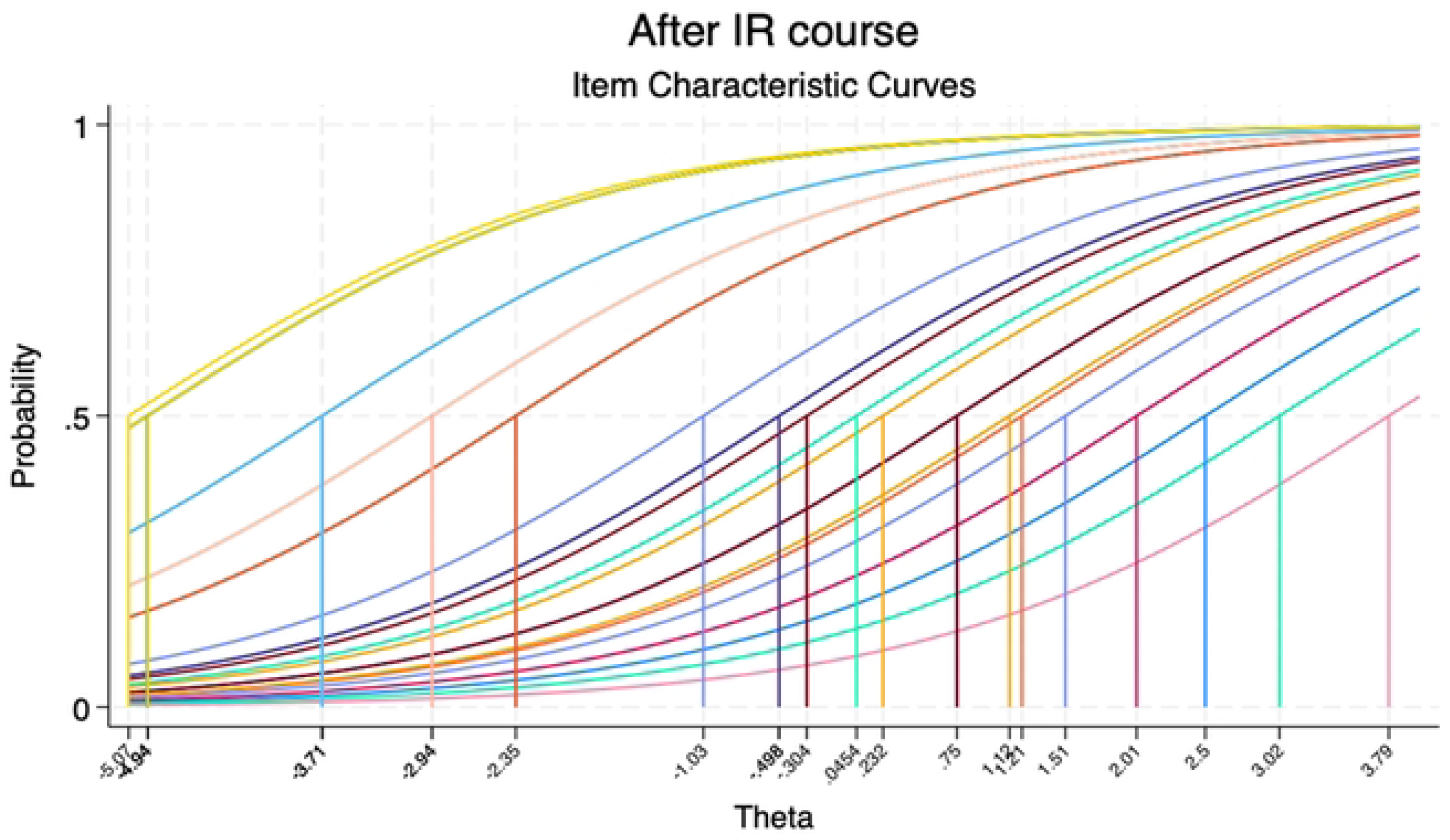
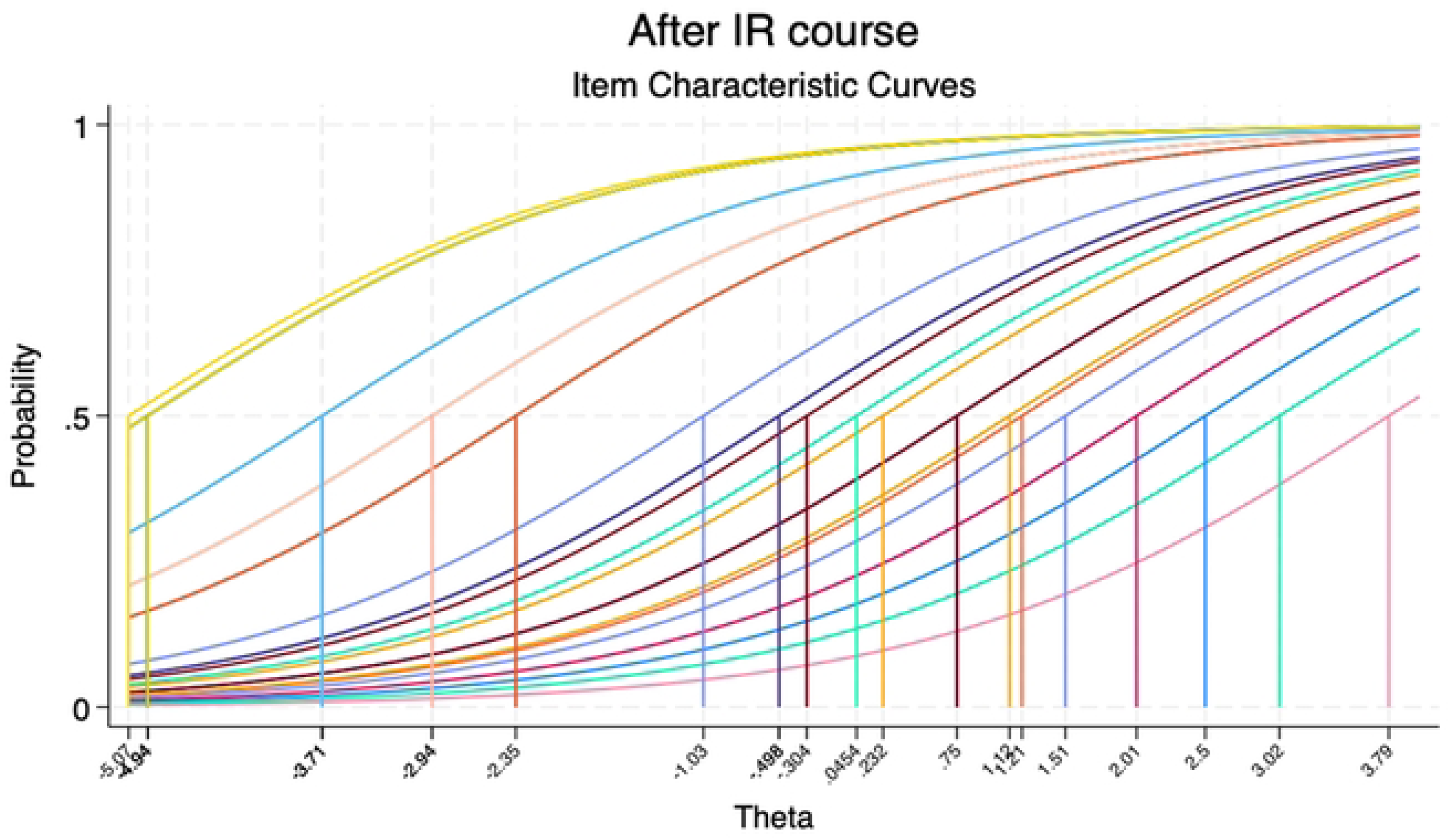
Pre- and post-Institute item characteristics for the objective assessment questionnaire assessing the difficulty of the questions. Fig 1 (a) Item Characteristic Curves (ICC) before the course Fig 2 Item Characteristic Curves (ICC) after the course

**Table 5.** Objective evaluation of Implementation Research knowledge.

| Period of IR Course | Before<br>n = 20 | After<br>n = 21 | P-value |
| --- | --- | --- | --- |
| **Theme 1: Working with stakeholders n (%)** | **Correct Responses** | **Correct Responses** |  |
| Stakeholders include any group or individual who can affect or is affected by the achievement of a specified objective. | 20 (100.0) | 21 (100.0) | N/A |
| The definition of a relevant stakeholder group typically begins with a broad review of health organizations in a geographical area. | 5 (25.0) | 4 (19.0) | 0.645 |
| Techniques (or approaches) for identifying stakeholders may include a multi-step (or multi-stage) process of conveying and working with different relevant groups to ensure representation, credibility, and legitimacy. | 20 (100.0) | 21 (100.0) | N/A |
| Some techniques for analyzing stakeholder interests and views of a specified objective may include brainstorming on the interests of different stakeholders and views on what can be done to satisfy those interests. | 20 (100.0) | 21 (100.0) | N/A |
| Power versus interest grids is an example of a technique for identifying and analyzing stakeholders. It typically categorizes stakeholders into two categories: players and subjects. | 1 (5.0) | 0 (0.0) | 0.300 |
| Ranking of stakeholders according to their importance to a specified objective may be necessary as part of stakeholder analysis. This ranking should be guided mainly by the stakeholder's experience. | 8 (40.0) | 5 (23.8) | 0.265 |
| Participatory research involves sharing decision-making power with stakeholders, especially at the community level in setting research agenda, conducting the research, and interpreting the findings and implementing actions as part of the research process. | 20 (100.0) | 21 (100.0) | N/A |
| **Theme 2: Scientific inquiry n (%)** |  |  |  |
| Implementation research outcomes (IRO) are useful for measuring the success or failure of a set of implementation activities. Examples of IROs include measures of mortality and morbidity. | 6 (30.0) | 12 (57.1) | 0.080 |
| IR prioritizes research on the systematic uptake of evidence-based knowledge and practice to improve services and population health. | 20 (100.0) | 20 (95.2) | 0.323 |
| One of the reasons for using theories, models, and frameworks in implementation research include testing the efficacy of new interventions in controlled settings. | 5 (25.0) | 12 (57.1) | 0.037 |
| Process models such as the Knowledge-to-Action Framework are mainly used for explaining what influences implementation outcomes. | 4 (20.0) | 7 (33.3) | 0.335 |
| Participatory research involves sharing decision-making power with stakeholders, especially at the community level, in setting research agenda, conducting the research, interpreting the findings, and implementing actions as part of the research process. | 20 (100.0) | 21 (100.0) | N/A |
| The Consolidated Framework for IR is an example of a determinant framework useful for understanding how contextual factors affect implementation outcomes. | 18 (94.7) | 20 (100.0) | 0.299 |
| Psychological theories such as the Theory of Planned Behavior are not useful in IR. | 18 (90.0) | 18 (90.0) | 1.000 |
| There is a tension between controlling conditions of the research study and ensuring external validity of the findings to other settings and IR prioritizes controlling the conditions of the research study. | 8 (40.0) | 11 (55.0) | 0.342 |
| Effectiveness-implementation hybrid design Type 3 prioritizes the use of quantitative methods to study the effect of clinical interventions on their related outcomes. | 5 (25.0) | 10 (50.0) | 0.102 |
| IR questions may be framed around understanding conditions that support the introduction of an evidence-based intervention into new settings. | 18 (94.7) | 20 (100.0) | 0.299 |
| IR questions involve testing the efficacy of interventions for improving clinical outcomes. | 9 (47.4) | 9 (47.4) | 1.000 |
| Approaches for measuring implementation fidelity include exit interviews, observations, or the use of checklists to observe adherence to the intervention protocol. | 19 (100.0) | 20 (100.0) | N/A |
| Measurement of implementation fidelity is useful at any stage of the implementation process. | 4 (21.1) | 6 (30.0) | 0.522 |
| **Theme 3: Implementation strategies n (%)** |  |  |  |
| Implementation strategies are techniques, activities or approaches used to enhance the adoption, implementation, and sustainability of evidence-supported interventions. | 20 (100.0) | 20 (100.0) | N/A |
| Identification of strategies for facilitating implementation of evidence-supported interventions and conduct of IR should be conducted independent of the stakeholder consultation process. | 15 (78.9) | 18 (90.0) | 0.339 |
| Compared to a comprehensive review, a rapid review may be restricted based on the breadth of the question(s) being explored. | 18 (94.7) | 19 (95.0) | 0.970 |
| The major recommended guidelines for measuring or reporting an implementation strategy are to specify the action and implementation outcomes targeted by the strategy. | 0 (0.0) | 2 (10.0) | 0.157 |
| Stand-alone strategies addressing a specific outcome at the individual or community level are more effective than multifaceted strategies that address multiple outcomes at multiple levels. | 12 (63.2) | 16 (80.0) | 0.243 |
| **Theme 4: Resources n (%)** |  |  |  |
| Implementation teams are required to effectively and efficiently support use of evidence-supported interventions in practice. | 20 (100.0) | 19 (95.0) | 0.311 |
| IR teams should include only researchers with expertise in quantitative and qualitative research methods. | 18 (90.0) | 17 (85.0) | 0.633 |
| Techniques (or approaches) for identifying stakeholders may include a multi-step (or multi-stage) process of conveying and working with different relevant groups to ensure representation, credibility, and legitimacy. | 20 (100.0) | 19 (95.0) | 0.311 |
| Some techniques for analyzing stakeholder interests and views of a specified objective may include brainstorming on the interests of different stakeholders and views on what can be done to satisfy those interests. | 20 (100.0) | 19 (95.0) | 0.311 |
| **Theme 5: Communication and advocacy n (%)** |  |  |  |
| Framing IR results to appeal to a specific disadvantaged group is more persuasive than framing the results to appeal to a broader stakeholder group. | 8 (40.0) | 8 (40.0) | 1.000 |
| Framing IR results to include numerical point estimates of effectiveness are more persuasive than using narratives and non-numerical recommendations. | 11 (55.0) | 16 (80.0) | 0.091 |
| Channels for disseminating implementation results to practitioners and policymakers include use of news media, social media, one-on-one meetings, and policy briefs. | 19 (95.0) | 18 (90.0) | 0.548 |
| Communicating what is lost by not using IR results is more effective than communication on what can be gained. | 8 (40.0) | 8 (40.0) | 1.000 |
| **Theme 6: Cross-cutting concepts n (%)** |  |  |  |
| Organizational readiness for change is defined as the extent to which organizational members are ready to implement organizational change e.g. adopt an evidence-supported intervention. | 20 (100.0) | 20 (100.0) | N/A |
| Conducting an organization readiness assessment prior to implementing an evidence-supported intervention helps to forecast and plan resources for implementation. | 19 (100.0) | 20 (100.0) | N/A |
| Organizational readiness for change can be assessed by reviewing history of prior implementation of evidence-supported interventions in different organizations. | 4 (20.0) | 0 (0.0) | 0.035 |
| Implementation climate is positively associated with implementation effectiveness and can be measured by three constructs: efficacy, valence, and motivation. | 4 (20.0) | 3 (15.0) | 0.677 |
| According to the Consolidated Framework for Implementation Research (CFIR), the outer setting of an implementing organization includes the culture of the organization and the prevailing implementation climate. | 5 (26.3) | 7 (35.0) | 0.557 |
| Clinical equipoise (lack of knowledge whether an intervention is better than its comparison) is generally used to justify the conduct of IR. | 10 (50.0) | 13 (65.0) | 0.337 |
| Challenges in operationalizing the process of informed consent in conducting IR may include challenges in identifying the actual research beneficiaries and participants. | 17 (85.0) | 17 (85.0) | 1.000 |
| **Summary Scores** |  |  |  |
| Percentage Total Score, mean (sd) | 65.35 (9.27) | 68.18 (8.87) | 0.324 |
| Percentage Total Score, mean (95% ci) | 65.35 (61.28; 69.41) | 68.18 (64.38; 71.97) | 0.324 |
| Total Score, mean (sd) | 25.80 (3.87) | 26.57 (5.66) | 0.615 |
| Total Score, mean (95% ci) | 25.80 (24.10; 27.50) | 26.57 (24.15; 28.99) | 0.615 |

Notes: ci=confidence interval; IR=implementation research; sd=standard deviation

## DISCUSSION

IR is widely recognized as a tool to bridge the gap between research and practice, ensure appropriate contextual adaptations, provide solutions to complex global health challenges, increase efficiency, capacity, and sustainability of implemented programs, and provide evidence-based data for global health practice and policymakers.[22] Despite a number of available trainings in IR[23–25], ranging from in-person or remote webinars to short courses and academic programs, many are inaccessible or fail to have a real impact on participant competencies and confidence in applying presented material in real-life settings. Additionally, a few programs are provided in the global health space[23] with a focus on equity issues.

SCGH SI provided a one-week intensive workshop that introduced IR in global health settings and effectively delivered training on the application of IR concepts and methods to address health inequities in global health research and practice. The design of SCGH SI and assessments focused on increasing competencies across various IR and confidence in the application of presented methodologies in real-life settings.

Competency-based training [12, 13] is recognized as a path to the effective application and training of global health practitioners to achieve impact at scale; however, many trainings fail to achieve an increase in competencies or achieve impact across important competency domains. For example, evaluation of an established brief virtual IS training program reported improvement across domains, particularly in areas of definition, background, and rationale, while the competencies related to design and analysis showed the lowest improvement.[26] Evaluation suggests lower confidence in the application of IR and suggests the need for more case-based learning. Many training courses also fail to apply relevant and validated assessments to evaluate changes in competency-based skill development during the training. In the context of SCGH SI, the pre-SI self-assessments showed that more than half of the participants had already attended some courses introducing IR methodology. Further, many participants expressed some knowledge and confidence in certain IR competencies, particularly in stakeholder engagement and basic IR theories. However, the pre-SI objective assessment revealed gaps in competencies across domains. The post-assessments and projects developed within the SI highlighted the importance of training in the application of IR strategies to real-world solutions as emphasized by other training courses.[23]

During SCGH SI, participants had the opportunity to apply newly acquired or enhanced skills in IR and equity to their projects and contexts and present their projects to peers and field expert trainers. This aspect of the SI – practical application of IR methodologies – was reflected in the self-assessed and objectively assessed increased knowledge and confidence in the utilization of IR concepts. Projects were developed by participants before attending the SI, often based in their geographical area and field. Given the residence and research fields, it is not surprising that most projects were set in LMICs, particularly in the African Region and Southeast Asia. The majority of topics focused on many priority areas in global health and health equity, and the focus of the Sustainable Development Goals (SDGs)[27], including maternal and child health, infectious diseases (particularly HIV), non-communicable diseases (cardiovascular issues, various types of cancer), mental health, and more, as well as crosscutting issues. Most participants in the SI were already established experts in their fields and were comfortable and confident in utilizing the methodology. Throughout SI discussions, participants expressed that they see the IR and equity framework as a path for addressing methodological and theoretical gaps in their research and practice, which they have not been able to fully address within their current field and existing methodologies. During the SI, the issues addressed and geopolitical areas indicate why these participants would be interested in developing competencies in IR methodologies with a particular focus on health equity.[4]

In their projects, most participants selected to utilize the Consolidated Framework for Implementation Research (CFIR), Exploration, Preparation, Intervention, Sustainment (EPIS), and Reach, Effectiveness, Adoption, Implementation, Maintenance (RE-AIM) frameworks. These were often in combination with other theoretical models and frameworks, including Health Beliefs and the Socioecological Model. Participants thus thoughtfully adapted and built upon their expertise and existing theories, models, and frameworks of their field to take advantage of IR and simultaneously advance IS.[4] While these theories, models, and frameworks were correctly applied, they may have been utilized because of the explicit coverage during the SI training and developed confidence in the application of these particular approaches. Additionally, participants expressed the need for further training and advanced methodologies that can be adapted and are suitable for their settings and contexts.

SI participants were also explicitly trained and required to incorporate equity considerations into their projects. Most approached it by examining their populations and vulnerable geopolitical settings as indicators of potential health inequities. As discussed, projects were set in LMICs, a vulnerable setting in its own right. However, participants uncovered vulnerabilities within LMICs, including marginalized communities, socioeconomically disadvantaged populations, remote and rural hard-to-access populations, people living with stigmatized conditions, and health system vulnerabilities. While literature is evolving around the use of IR in global health and LMIC settings, critical gaps remain in how to specifically tackle various global health challenges, and few studies explicitly examine health inequities. However, it has become clear that IR must evolve to produce contextually grounded methodologies suitable for LMIC and resource-limited settings.[4] Some proposed approaches to address inequities through IR include explicating and monitoring outcomes for vulnerable groups, participatory approaches involving vulnerable groups in defining and implementing the IR agenda, and strategies addressing social and structural determinants of health – consistent with approaches suggested in the literature.[2, 5]

This paper further addresses the gaps that exist in the objective assessment of the impact of IR trainings[23], particularly in the context of short-term training, by utilizing and validating tools for competency-based assessments of IR training explicitly developed for the low- and middle-income country settings [15]. The SCGH SI created a collaborative and enriching environment with diverse topics and geographic focus, facilitating networking opportunities and potential partnerships for future programs. This aspect of SCGH SI training is reflected by significant increases in scores across domains such as project co-design and stakeholder engagement. The positive collaborative setting during SCGH SI reflected high satisfaction with SI’s structure, content, and delivery. Participants emphasized the need for continued collaboration between SCGH SI trainers and peers. Furthermore, IR methodologies and applications are rapidly developing, underscoring the need for continuous IR training development and enhancement of new areas and aspects. Consideration of further competencies will be identified as critical in the effective application of IR. In addition to the need to provide training beyond basic concepts and short-term delivery, further resources and training may be developed to maintain competencies gained. While SCGH SI was provided in-person, accessibility of online and free-source resources for further impact is necessary to achieve impact on a global scale. Future assessments may provide valid comparisons of the impact of online versus in-person delivery.

### Strengths and Weaknesses

To our knowledge, this is one of the first studies that rigorously evaluates IR training in global health and equity, utilizing competency-based assessment adapted for LMIC settings, and providing further insight into an effective design of such training. This study was not without limitations. The assessments utilized have not shown a correlation between self-assessed and objectively assessed knowledge of IR concepts. A paired analysis was not feasible given an error in capturing unique identifiers. For future assessments, we will implement an approach to assign personalized links which will allow a paired analysis and a more accurate assessment at an individual level, as well as association with personal and professional characteristics and prior experience with IR concepts.

This paper aims to provide insights into the impact of competency-based IR training programs for researchers and practitioners, and how these programs can be designed to address advanced competencies in IR, including tackling health equity [23, 26]. The findings from this paper also contribute to the methods and tools for competency-based assessment of IR training and will be used to improve the SCGH SI and similar training programs in global health settings. In the future, the SCGH SI aims to increase its impact and evaluation including (1) expanding the reach to practitioners going beyond academia-based researchers, offering the SI in other geopolitical locations, and online/hybrid delivery; (2) offering more personalized training based on the participant pre-SI competency level by offering simultaneous beginner and intermediate/advanced blended course with adaptations and level-based activities or separate beginner and advanced level trainings, and expanding competencies; (3) developing a rigorous plan for continuous engagement with trainees post-SI; (4) training the participants to lead SI in their locations; and (5) continuing to refine assessments and assessing long-term impact of SCGH SI, e.g., in the area of competency retainment, funding, and publications.

## CONCLUSIONS

Implementation science is uniquely positioned to target inequity-reducing strategies and fast-track progress toward global health and meeting the SDGs. The provision of an intensive workshop focused on IS and equity with intentional hands-on practice and support from mentors and peers demonstrates the potential to efficiently train implementers of global health solutions across various settings and topical areas. Continued refining of assessment tools and evaluating the long-term impact of training will further contribute to the effective design and implementation of research training in global health.

## Acknowledgments

We acknowledge the support of the UAB School of Public Health and especially our Summer Institute participants.

## Ethical Approval

Ethical approval for this study has been obtained from the Institutional Review Board at the University of Alabama at Birmingham, protocol number IRB-300013258.

## Informed Consent and Potential Risks

Not applicable.

## Consent to publication

Not applicable.

## Code availability

Not applicable.

## Author’s contribution

OA conceptualized the institute, evaluation design, and manuscript. AH and KO wrote the initial draft. AH conducted data collection. KO conducted the data analysis. AH, KO, and OA conducted the result interpretation. AH, KO, CD, ET, and OA revised the subsequent draft of the manuscript. OA is the guarantor for this paper.

## Data availability

The data that supports the findings of this study will be available from the corresponding author upon reasonable request. The final dataset will include assessment data and self-reported demographics. Upon completion of the study analysis, the final dataset will be stripped of identifiers prior to release for sharing.

## REFERENCES

1. Peters DH, Adam T, Alonge O, Agyepong IA, Tran N. Implementation research: what it is and how to do it. Bmj. 2013;347:f6753. Epub 20131120. doi: 10.1136/bmj.f6753. PubMed PMID: 24259324.

2. Alonge O. How to leverage implementation research for equity in global health. Glob Health Res Policy. 2024;9(1):43. Epub 20241017. doi: 10.1186/s41256-024-00388-5. PubMed PMID: 39420430; PubMed Central PMCID: PMCPMC11484107.

3. Braveman P, Arkin E, Orleans T, Proctor D, Plough A, Robert Wood Johnson Foundation ib, et al. What is health equity and what difference does a definition make? Princeton, NJ: Robert Wood Johnson Foundation; 2017.

4. Alonge O, Brooks MB. Implementation science derived from low- and middle-income countries is essential for advancing global health. BMC Glob Public Health. 2025;3(1):84. Epub 20250922. doi: 10.1186/s44263-025-00206-1. PubMed PMID: 40983963; PubMed Central PMCID: PMCPMC12452020.

5. Alonge O. Using causal models and theories to achieve equitable implementation science in global health. BMC Glob Public Health. 2025;3(1):85. Epub 20251001. doi: 10.1186/s44263-025-00203-4. PubMed PMID: 41029768; PubMed Central PMCID: PMCPMC12486490.

6. Woodward EN, Singh RS, Ndebele-Ngwenya P, Melgar Castillo A, Dickson KS, Kirchner JE. A more practical guide to incorporating health equity domains in implementation determinant frameworks. Implement Sci Commun. 2021;2(1):61. Epub 20210605. doi: 10.1186/s43058-021-00146-5. PubMed PMID: 34090524; PubMed Central PMCID: PMCPMC8178842.

7. Welten VM, Dabekaussen K, Melnitchouk N. Global Health 101. Clin Colon Rectal Surg. 2022;35(5):355–61. Epub 20220913. doi: 10.1055/s-0042-1746184. PubMed PMID: 36111085; PubMed Central PMCID: PMCPMC9470279.

8. Koplan JP, Bond TC, Merson MH, Reddy KS, Rodriguez MH, Sewankambo NK, et al. Towards a common definition of global health. Lancet. 2009;373(9679):1993–5. Epub 20090601. doi: 10.1016/s0140-6736(09)60332-9. PubMed PMID: 19493564; PubMed Central PMCID: PMCPMC9905260.

9. Chambers DA, Emmons KM. Navigating the field of implementation science towards maturity: challenges and opportunities. Implement Sci. 2024;19(1):26. Epub 20240313. doi: 10.1186/s13012-024-01352-0. PubMed PMID: 38481286; PubMed Central PMCID: PMCPMC10936041.

10. Brownson RC, Cabassa LJ, Drake BF, Shelton RC. Closing the gap: advancing implementation science through training and capacity building. Implement Sci. 2024;19(1):46. Epub 20240703. doi: 10.1186/s13012-024-01371-x. PubMed PMID: 38961482; PubMed Central PMCID: PMCPMC11223366.

11. Penkunas MJ, Ross B, Scott CP, Thorson A, Baron LF, Rebai WK, et al. Barriers to Applying Knowledge Gained Through an Implementation Research Massive Open Online Course: An Explanatory Qualitative Study. Inquiry. 2024;61:469580241284916. doi: 10.1177/00469580241284916. PubMed PMID: 39548770; PubMed Central PMCID: PMCPMC11569480.

12. Schleiff MJ, Mburugu PM, Cape J, Mwenesi R, Sirili N, Tackett S, et al. Training Curriculum, Skills, and Competencies for Global Health Leaders: Good Practices and Lessons Learned. Ann Glob Health. 2021;87(1):64. Epub 20210712. doi: 10.5334/aogh.3212. PubMed PMID: 34307067; PubMed Central PMCID: PMCPMC8284497.

13. Hansoti B, Hahn E, Dolive C, Akridge A, Atwell M, Mishra A, et al. Training Global Health Leaders: A Critical Review of Competency Gaps. Ann Glob Health. 2021;87(1):65. Epub 20210712. doi: 10.5334/aogh.3260. PubMed PMID: 34307068; PubMed Central PMCID: PMCPMC8284503.

14. Alonge O, Rao A, Kalbarczyk A, Maher D, Gonzalez Marulanda ER, Sarker M, et al. Developing a framework of core competencies in implementation research for low/middle-income countries. BMJ Glob Health. 2019;4(5):e001747. Epub 20190903. doi: 10.1136/bmjgh-2019-001747. PubMed PMID: 31544004; PubMed Central PMCID: PMCPMC6730585.

15. Alonge O, Rao A, Kalbarczyk A, Ibisomi L, Dako-Gyeke P, Mahendradhata Y, et al. Multimethods study to develop tools for competency-based assessments of implementation research training programmes in low and middle-income countries. BMJ Open. 2024;14(7):e082250. Epub 20240716. doi: 10.1136/bmjopen-2023-082250. PubMed PMID: 39013650; PubMed Central PMCID: PMCPMC11288145.

16. Eslava-Schmalbach J, Garzón-Orjuela N, Elias V, Reveiz L, Tran N, Langlois EV. Conceptual framework of equity-focused implementation research for health programs (EquIR). International Journal for Equity in Health. 2019;18(1):80. doi: 10.1186/s12939-019-0984-4.

17. Kiger ME, Varpio L. Thematic analysis of qualitative data: AMEE Guide No. 131. Med Teach. 2020;42(8):846–54. Epub 20200501. doi: 10.1080/0142159x.2020.1755030. PubMed PMID: 32356468.

18. Braun V, Clarke V. What can “thematic analysis” offer health and wellbeing researchers? Int J Qual Stud Health Well-being. 2014;9:26152. Epub 20141016. doi: 10.3402/qhw.v9.26152. PubMed PMID: 25326092; PubMed Central PMCID: PMCPMC4201665.

19. Consolidated Framework for Implementation Research (CFIR): CFIR Research Team-Center for Clinical Management Research; [cited 2025]. Available from: https://cfirguide.org/.

20. Aarons GA, Hurlburt M, Horwitz SM. Advancing a conceptual model of evidence-based practice implementation in public service sectors. Adm Policy Ment Health. 2011;38(1):4–23. doi: 10.1007/s10488-010-0327-7. PubMed PMID: 21197565; PubMed Central PMCID: PMCPMC3025110.

21. Holtrop JS, Estabrooks PA, Gaglio B, Harden SM, Kessler RS, King DK, et al. Understanding and applying the RE-AIM framework: Clarifications and resources. J Clin Transl Sci. 2021;5(1):e126. Epub 20210514. doi: 10.1017/cts.2021.789. PubMed PMID: 34367671; PubMed Central PMCID: PMCPMC8327549.

22. Theobald S, Brandes N, Gyapong M, El-Saharty S, Proctor E, Diaz T, et al. Implementation research: new imperatives and opportunities in global health. Lancet. 2018;392(10160):2214–28. Epub 20181009. doi: 10.1016/s0140-6736(18)32205-0. PubMed PMID: 30314860.

23. Davis R, D’Lima D. Building capacity in dissemination and implementation science: a systematic review of the academic literature on teaching and training initiatives. Implement Sci. 2020;15(1):97. Epub 20201030. doi: 10.1186/s13012-020-01051-6. PubMed PMID: 33126909; PubMed Central PMCID: PMCPMC7597006.

24. Baumann AA, Carothers BJ, Landsverk J, Kryzer E, Aarons GA, Brownson RC, et al. Evaluation of the Implementation Research Institute: Trainees’ Publications and Grant Productivity. Adm Policy Ment Health. 2020;47(2):254–64. doi: 10.1007/s10488-019-00977-4. PubMed PMID: 31667667; PubMed Central PMCID: PMCPMC7285898.

25. Means AR, Phillips DE, Lurton G, Njoroge A, Furere SM, Liu R, et al. The role of implementation science training in global health: from the perspective of graduates of the field’s first dedicated doctoral program. Glob Health Action. 2016;9:31899. Epub 20161114. doi: 10.3402/gha.v9.31899. PubMed PMID: 27846928; PubMed Central PMCID: PMCPMC5110555.

26. Van Pelt AE, Bonafide CP, Rendle KA, Wolk C, Shea JA, Bettencourt A, et al. Evaluation of a brief virtual implementation science training program: the Penn Implementation Science Institute. Implement Sci Commun. 2023;4(1):131. Epub 20231106. doi: 10.1186/s43058-023-00512-5. PubMed PMID: 37932840; PubMed Central PMCID: PMCPMC10626776.

27. Rasanathan K, Diaz T. Research on health equity in the SDG era: the urgent need for greater focus on implementation. International Journal for Equity in Health. 2016;15(1):202. doi: 10.1186/s12939-016-0493-7.

